# Fc-dependent functional activity of ChAdOx1-S and CoronaVac vaccine-induced antibodies to the SARS-CoV-2 spike protein

**DOI:** 10.1101/2023.10.25.23297503

**Authors:** Alexander W. Harris, Liriye Kurtovic, Jeane Nogueira, Isabel Bouzas, D. Herbert Opi, Bruce D. Wines, P. Mark Hogarth, Pantelis Poumbourios, Heidi E. Drummer, Clarissa Valim, Luís Cristóvão Porto, James G. Beeson

## Abstract

Ongoing severe acute respiratory syndrome coronavirus 2 (SARS-CoV-2) transmission and COVID-19 disease severity is influenced by immunity acquired by natural exposure and/or vaccination, whereby most vaccines are formulated on the Ancestral strain. However, population-level immunity is complicated by the emergence of variants of concern (VOCs), such as Omicron that is the dominant variant currently in circulation. Antibody Fc-dependent effector functions are being increasingly recognised as important mediators in immunity, especially against VOCs. However, induction of these functions in populations with diverse infection and/or vaccination histories, remains poorly defined. Here, we evaluated Fc-dependent functional antibodies following vaccination with two widely used vaccines: AstraZeneca (AZ; ChAdOx1-S) and Sinovac (SV). We quantified FcγR-binding and C1q-fixing antibodies against Ancestral and variant spike (S) proteins in Brazilian adults vaccinated with AZ or SV (n=222), some of which were previously exposed to SARS-CoV-2. AZ induced greater FcγR-binding responses to Ancestral S than the SV vaccine. Previously exposed individuals had significantly greater vaccine-induced responses compared to their naïve counterparts, with notably high C1q-fixation levels, irrespective of vaccine type. FcγR-binding was highest among AZ vaccinated individuals with a prior exposure, and these responses were well retained against the Omicron S protein. Overall, these findings contribute to our understanding of vaccine-induced immunity and its effectiveness against evolving variants.

## Introduction

The emergence of severe acute respiratory syndrome coronavirus 2 (SARS-CoV-2) in December 2019 has caused more than 770 million cases of COVID-19 and nearly 7 million deaths globally by August 2023 (1). The development and deployment of efficacious vaccines formulated on the original Ancestral (Hu-1) strain has helped reduce the global burden of COVID-19, however, this progress is threatened by the emergence of variants of concern (VOCs). Ongoing SARS-CoV-2 transmission and COVID-19 disease in populations is influenced by heterologous immunity provided by partial vaccine coverage and acquired from natural infections. A large proportion of the global population remains unvaccinated but may have naturally acquired immunity following exposure to SARS-CoV-2. Different vaccines used globally may elicit different antibody response profiles, and the magnitude and protective functions of vaccine-induced antibodies are also affected by acquired immunity from prior SARS-CoV-2 infections. Greater knowledge of the specificity and functions of vaccines used globally is needed to define vaccination regimens, optimal vaccination coverage, understand the impact of VOCs and inform next-generation vaccines.

Disease burden and mortality rates were particularly high in Brazil, where the Oxford-AstraZeneca (ChAdOx1-S; AZ) and Sinovac Biotech CoronaVac (SV) vaccines were approved for emergency use in early 2021 (2). These vaccines were used extensively in Brazil (3). Globally, these vaccines are being deployed at mass-scale with over 1.5 billion doses of AZ and 1.8 billion doses of SV delivered world-wide in 2021 (4), and they continue to be used as part of COVID-19 vaccine access programs in Africa, such as COVAX (5).

The AZ vaccine is a viral vector-based vaccine that uses an adenovirus derived from chimpanzees to deliver coding sequences of the SARS-CoV-2 spike (S) protein involved in viral attachment and entry into host cells. Conversely, the SV vaccine uses inactivated virus containing the full range of proteins, including the S protein. A retrospective cohort study of over 60 million vaccinated Brazilians concluded that AZ and SV were associated with a 78.1% and 53.2% reduced risk of symptomatic illness, and a 92.3% and 73.7% reduced risk of death, respectively (6). However, a deeper understanding of the protective immune profiles induced by these vaccines is necessary to inform optimal booster policy and understand the retention of protection against VOCs.

The generation of SARS-CoV-2 specific immunoglobulin G (IgG) correlates strongly with vaccine efficacy (7) and protection from severe disease (8, 9). These antibodies can neutralise SARS-CoV-2 virions primarily by binding to the receptor binding domain (RBD) of the S protein to prevent viral entry into host cells via angiotensin-converting enzyme 2 (ACE-2) (10). The protective association of neutralising antibodies against SARS-CoV-2 infection and disease has been well documented (7, 9, 11). However, studies have reported greatly reduced neutralising titres in AZ (12) and SV (13, 14) vaccinated individuals against the Omicron (BA.1/B.1.1.529) VOC, which has fifteen mutations in the RBD compared to the ancestral vaccine strain (15), leading to increased breakthrough infections. However, despite the loss of antibody neutralising activity against Omicron and other VOCs, the vaccines still confer significant efficacy suggesting roles for other immune mechanisms in immunity.

An immunologic mechanism that is being increasingly recognised in immunity against SARS-CoV-2 and other pathogens is the role of antibodies engaging immune cells and the complement system via the fragment crystallisable region (Fc). These Fc-dependent effector functions have previously been identified as correlates of protection against other viruses such as influenza (16), HIV (17, 18), and Ebola (19). These functions can also be mediated by antibodies against epitopes outside the RBD (20). While less studied than virus neutralisation, Fc-dependent effector functions likely contribute to protection against the effects of SARS-CoV-2 infection and may be of particular importance in protection against emerging VOCs with RBD mutations where neutralisation potency is diminished.

IgG bound to antigens can engage Fc gamma receptor 3 (FcγRIII), which is highly expressed on natural killer (NK) cells and mediates direct killing of virally infected cells via antibody-dependent cellular cytotoxicity (ADCC) (21). Antibodies can also engage FcγRI, FcγRIIa, and FcγRIII and FcγRIIIb, expressed on phagocytic cells (e.g. macrophages, monocytes, and neutrophils) and promote antibody-dependent cellular phagocytosis (ADCP), to boost clearance of virions (22). FcγRI is a high-affinity receptor that readily binds IgG, whereas FcγRIIa and FcγRIII are low-affinity receptors which require multivalent binding by multiple IgG molecules for activation (23). These Fc-effector functions have been shown to improve survival in SARS-CoV-2 challenge mouse models using Fc engineered monoclonal antibodies (mAbs) and FcγR deletion (24–26) studies and observational studies of hospitalised patients (27). Antibody dependent complement deposition (ADCD) contributes to host immune defence by promoting enhanced antibody neutralisation, inflammation, direct lysis, and promoting phagocytosis of pathogens through binding to complement receptors (28). IgG complexes and pentameric IgM can both fix complement component C1q, activating the classical pathway of the complement system. While complement activation likely contributes to optimal protection against SARS-CoV-2, appropriate regulation is essential as elevated levels of downstream complement factors have been associated with severe disease and death (29, 30).

Few studies have evaluated Fc-functional antibody responses induced by vaccination with AZ or SV vaccines, especially in large cohorts. Greater knowledge is important given the extensive use of these vaccines globally and the continuing COVID-19 pandemic. An early phase 1/2 trial found that AZ vaccination induced greater ADCC, ADCP, and ADCD functions compared to unvaccinated COVID-19 patients with mild disease (31). Detectable levels of anti-S antibodies with ADCP activity have been demonstrated following SV vaccination, however, responses were less than that of convalescent individuals (32). Encouragingly, retention of FcγRIIa- and FcγRIII-binding following SV vaccination has been reported between the Ancestral and Omicron S proteins. However, reactivity was not maintained to the Omicron RBD (33), suggesting a preservation of Fc-dependent functional activity largely at non-RBD epitopes, albeit in a small sample size (n=13). Additionally, previous exposure to SARS-CoV-2 has been shown to boost anti-S IgG following AZ (34) or SV (35) vaccination, but it remains unknown how this impacts antibody Fc-functional activity.

Here, we investigated Fc-dependent functional activities of antibodies against the SARS-CoV-2 S protein in serum from a cohort of Brazilian adults that received the AZ or SV vaccines, and unvaccinated individuals with a mild-moderate SARS-CoV-2 infection. We determined the effect of previous SARS-CoV-2 infection on vaccine-induced antibody responses and evaluated Fc-dependent functional activity to the Delta (B.1.617.2) and Omicron VOC S proteins compared to the ancestral vaccine strain.

## Methods

### Study Population and Ethics Approval

Brazilian adults attending the health centre of Rio de Janeiro State University (UERJ) with a suspected SARS-CoV-2 infection or to receive a COVID-19 vaccine were invited to participate in the present study **(Table 1)**. The UERJ health centre provides secondary and specialised care for the Rio de Janeiro population and was a designated COVID-19 diagnostic site for personnel working in the Brazilian public health system during 2020 (36). It was later engaged in SARS-CoV-2 vaccination for students, employees, and the general population. Serum samples were collected from participants who received two doses of AstraZeneca (AZ) or Sinovac (SV) vaccines at a median of 34 days post-vaccination (n=222). Vaccine doses were administered at days 0 and 90 for AZ and at days 0 and 28 for SV. Of vaccinated individuals, 46.8% (n=104) had a known prior SARS-CoV-2 infection that occurred between 1/03/2020 and 1/01/2021. Serum samples were also collected from unvaccinated participants with a PCR confirmed mild-moderate SARS-CoV-2 infection between 1/01/2020 and 31/08/2020 at a median of 67 days post infection (n=200). Some individuals were included in both cohorts (n=20). Note that all infections recorded in this study occurred when the Ancestral strain was the dominant variant in circulation in Brazil. Participants provided written informed consent, and ethics approval was obtained by: National Commission on Research Ethics of UERJ, Brazil (CAAE: 30135320.0.0000.5259); Alfred Human Research and Ethics Committee, Australia (approval number: 307/22).

**Table 1.** Detailed cohort demographics.

|  | AstraZeneca Naïve (AZ-N) | AstraZeneca Exposed (AZ-E) | Sinovac Naïve (SV-N) | Sinovac Exposed (SV-E) | Natural Infection (I) | P |
| --- | --- | --- | --- | --- | --- | --- |
| n | 55 | 32 | 63 | 72 | 200 |  |
| Sex (female) | 38 | 24 | 48 | 58 | 143 | 0.9507* |
| Age (years) | 63<sup>a</sup> (61-69) | 62<sup>a</sup> (54-64) | 50<sup>b</sup> (40-59) | 45<sup>b</sup> (33-57) | 44<sup>b</sup> (36-51) | <0.05<sup>#</sup> |
| Sample date (range) | 19/5/21-10/9/21 | 25/5/21-10/9/21 | 22/3/21-20/8/21 | 24/3/21-12/8/21 | 11/5/20-9/2/21 |  |
| Days post second vaccination dose/primary infection | 31<sup>a</sup> (29-34) | 32<sup>a</sup> (29-35) | 34<sup>a</sup> (30-35) | 35<sup>a</sup> (30-37) | 67<sup>b</sup> (55-111) | <0.0001<sup>#</sup> |
| Days post previous exposure | - | 350 (207-443) | - | 354 (244-388) | - | 0.2604<sup>†</sup> |

Data presented as median and IQR unless specified otherwise. Values in a row without a common superscript letter differ significantly (P<0.05).
\*= $\chi^2$ , <sup>#</sup>=Kruskal-Wallis test followed by a Dunn's multiple comparisons test, and <sup>†</sup>= Mann-Whitney U test.

### SARS-CoV-2 Antigens

Antibody responses were measured to Ancestral S trimer (SPN-C52H4; SPN-C52H9), biotinylated Ancestral S trimer (SPN-C82E9), Delta B.1.617.2 S trimer (SPN-C52He), biotinylated Delta B.1.617.2 S trimer (SPN-C82Ec), Omicron BA.1/B.1.1.529 S trimer (SPN-C52Hz), and biotinylated Omicron BA.1/B.1.1.529 S trimer (SPN-C82Ee). Proteins were recombinantly expressed using the HEK293 expression system. Protein content and purity were validated by the manufacturer and in house using SDS-PAGE **(Figure S1)**. All SARS-CoV-2 antigens were commercially purchased from ACROBiosystems, USA.

### Antibody Detection by Enzyme-Linked Immunosorbent Assay (ELISA)

To detect antigen-specific antibodies (IgG, IgM, and IgA), we coated 384-well Spectra plates (PerkinElmer, USA) with Ancestral, Delta, or Omicron S trimer at 0.5 µg/ml in phosphate buffered saline (PBS) and incubated overnight at 4°C. The plates were blocked with 0.1% casein (Sigma-Aldrich, USA) in PBS (w/v) and serum samples were tested at 1/2000 (IgG), 1/800 (IgM), or 1/400 (IgA) dilution using the JANUS automated workstation (PerkinElmer) as previously validated (37). Isotype-specific antibodies were detected using goat anti-human IgG (1/1000; Invitrogen, USA), IgM (1/2500; Sigma-Aldrich), or IgA (1/5000; Abcam, UK) conjugated to horseradish peroxidase (HRP). The plates were then incubated with 3,3’, 5,5’ tetramethylbenzidine (TMB) substrate (Thermo Fisher Scientific, USA) and shielded from light. The colour-changing reaction was stopped using 1% H_2_SO_4_ in dH2O (v/v) and optical density (OD) was immediately read at 450nm using a Multiskan GO plate reader (Thermo Fisher Scientific). Serum samples and all reagents were prepared using 0.1% casein in PBS (w/v) unless specified otherwise. Note that for all plate-based assays, prior to addition of any reagent or serum, plates were washed thrice in a microplate washer (Millennium Science, AUS) using 0.05% Tween in PBS (v/v) unless specified otherwise.

### FcγR-binding Assays

FcγR-binding assays were conducted to quantify antibody binding to FcγRI, FcγRIIa, and FcγRIII as previously described (38–40). Briefly, 384-well Spectra plates were coated with Ancestral, Delta, or Omicron S trimer at 0.5 µg/ml in PBS and incubated overnight at 4°C. Plates were blocked with 1% bovine serum albumin (BSA; Sigma-Aldrich) in PBS (w/v) and serum samples were tested at 1/8000 (FcγRI) or 1/200 (FcγRIIa and FcγRIII) dilution using the JANUS automated workstation. Plates were then incubated with biotinylated human FcγRI (Sino Biological, CN) at 0.125 µg/ml, FcγRIIa-H131 dimer (40) at 0.2 µg/ml, or FcγRIII-V158 dimer (40) at 0.1 µg/ml, followed by the addition of streptavidin conjugated HRP (Invitrogen) at 1/10,000 dilution. Plates were then incubated with TMB, followed by 1% H_2_SO_4_ in dH2O (v/v) and absorbance was immediately read at 450 nm. Serum samples and all reagents were prepared in 1% BSA in PBS (w/v) unless specified otherwise.

To confirm that FcγR-binding correlated with cellular effector functions, we evaluated the ability of serum from vaccinated individuals to activate cells expressing FcγRIIa or FcγRIII *in vitro* as previously described (41). Serum samples were randomly selected from the upper (n=20) and lower (n=20) tertiles of FcγRIIa- and FcγRIII-binding responders. Briefly, A549 cells expressing the full-length SARS-CoV-2 spike protein (A549S) were allowed to adhere to wells of 96 well opaque plates (Corning, USA) and serum samples were added in duplicate at a dilution of 1/200 in RPMI containing 5% FCS, 2mM glutamine, 55 µM 2-merceptoethanol, 100units/ml Penicillin, and 100 µg/ml Streptomycin to allow for antibody opsonisation. Plates were washed thrice with RPMI media before IIA1.6 reporter cells with a NF-κB-nanoluciferase FcγRIIa H131 or NF-κB-nanoluciferase FcγRIII V158 construct were added. Following incubation, cells were lysed by the addition of Nano-Glo luciferase assay substrate at a dilution of 1/1000 in 10 mM Tris pH 7.4, containing 5 mM EDTA, 0.5 mM DTT, and 0.2% Igepal CA-630 (Sigma Aldrich). The induction of nano-luciferase expression was determined by measuring the relative luminescence units (RLU) of each well for one second/well using a Clariostar Optima plate reader (BMG Labtech, USA).

### Complement-Fixation Assay

Assays to quantify antibody binding to the hexameric complement component C1q, which is required to initiate the classical component pathway, were conducted as previously described (42). Briefly, 384-well Spectra plates were coated with 5 µg/ml of avidin (Jomar Life Research, AUS) in PBS. Plates were blocked with 0.1% casein in PBS (w/v) and biotinylated S protein was added at 1 µg/ml in PBS. Serum samples were tested at 1/100 dilution using the JANUS automated workstation, followed by the addition of purified human C1q (MilliporeSigma, USA) at 10 µg/ml. Plates were incubated with rabbit anti-C1q IgG (42, 43) at 1/2000 dilution, followed by goat anti-rabbit IgG HRP (MilliporeSigma) at 1/2000 dilution. Plates were then incubated with TMB, followed by 1% H_2_SO_4_ in dH2O (v/v) and absorbance was immediately read at 450 nm. All serum and reagent dilutions were performed using 0.1% casein in PBS (w/v).

To confirm that C1q-fixation led to the activation of downstream complement proteins, we evaluated the ability of serum from vaccinated individuals to fix the terminal complement component C5b-C9 that forms the membrane attack complex. Serum samples were randomly selected from the upper (n=20) and lower (n=20) tertiles of IgG responders. Plates were prepared as above, but human serum from pre-pandemic COVID-19 negative donors was used as a source of complement at 1/10 dilution and C5b-C9-fixation was detected using rabbit-anti-C5b-C9 IgG (Millipore) and goat anti-rabbit IgG HRP antibodies at 1/1000 dilution.

### Statistical Analysis

Raw data were corrected for background reactivity using no-serum controls. Positive control samples were used to standardise for plate-to-plate variability and negative control samples from pre-pandemic Melbourne donors were used to interpret seropositivity, which was defined as the mean + 2 standard deviations of the Melbourne donors (n=28). The following non-parametric statistical analyses were performed using GraphPad Prism 8 and Stata. Mann-Whitney U test to compare two independent groups. Kruskal Wallis test or a Friedman test, followed by a Dunn’s multiple comparisons test, to compare unpaired or paired data (respectively) in more than two groups. Spearman’s rank correlations to compare the relationship between antibody magnitude and functional responses. P<0.05 was considered statistically significant. Formal adjustment of p-values was not performed for multiple comparisons. Instead, the interpretation of differences between groups considered the p-value, the effect size, and the overall coherency of the findings.

## Results

### Fc-mediated antibody functions induced by AZ and SV vaccination in SARS-CoV-2 naïve individuals

We evaluated antibodies among participants who received two doses of the AZ or SV vaccines and were naïve to natural SARS-CoV-2 infection (AZ-N, n=55; SV-N, n=63), and unvaccinated individuals with a known SARS-CoV-2 infection (I, n=200). Serum samples collected post-vaccination (median [IQR] days: AZ, 31 [29-34]; SV, 34 [30-35]) or post-infection (median [IQR] days: 67 [55-111]) were tested in immunoassays against the Ancestral SARS-CoV-2 S protein (**Tabe 1**).

IgG magnitude to the Ancestral S protein was significantly higher in participants vaccinated with AZ compared to SV (P=0.0158; **Figure 1A**). IgG magnitude was similar between SV vaccinated individuals and individuals who had experienced a prior SARS-CoV-2 infection (P>0.9999). Both AZ and SV vaccines effectively induced antibodies able to engage FcγRI, FcγRIIa, and FcγRIII, as did antibodies from natural infection; however, the magnitude of activity varied substantially among individuals. Antibodies from AZ vaccinated individuals had greater activity than SV vaccinated individuals for all FcγRs (**Figure 1B-D**). The difference in response between vaccine groups was somewhat more prominent for FcγR-binding activity than for IgG, which might be relevant to differences in protective efficacy between the vaccines (median OD difference for AZ and SV groups: IgG, 0.39; FcγRI, 0.54; FcγRIIa, 0.52; FcγRIII, 0.59). IgG magnitude generally correlated strongly with FcγRI-, FcγRIIa-, and FcγRIII-binding across all groups (**Figure S2**).

**Figure 1.**
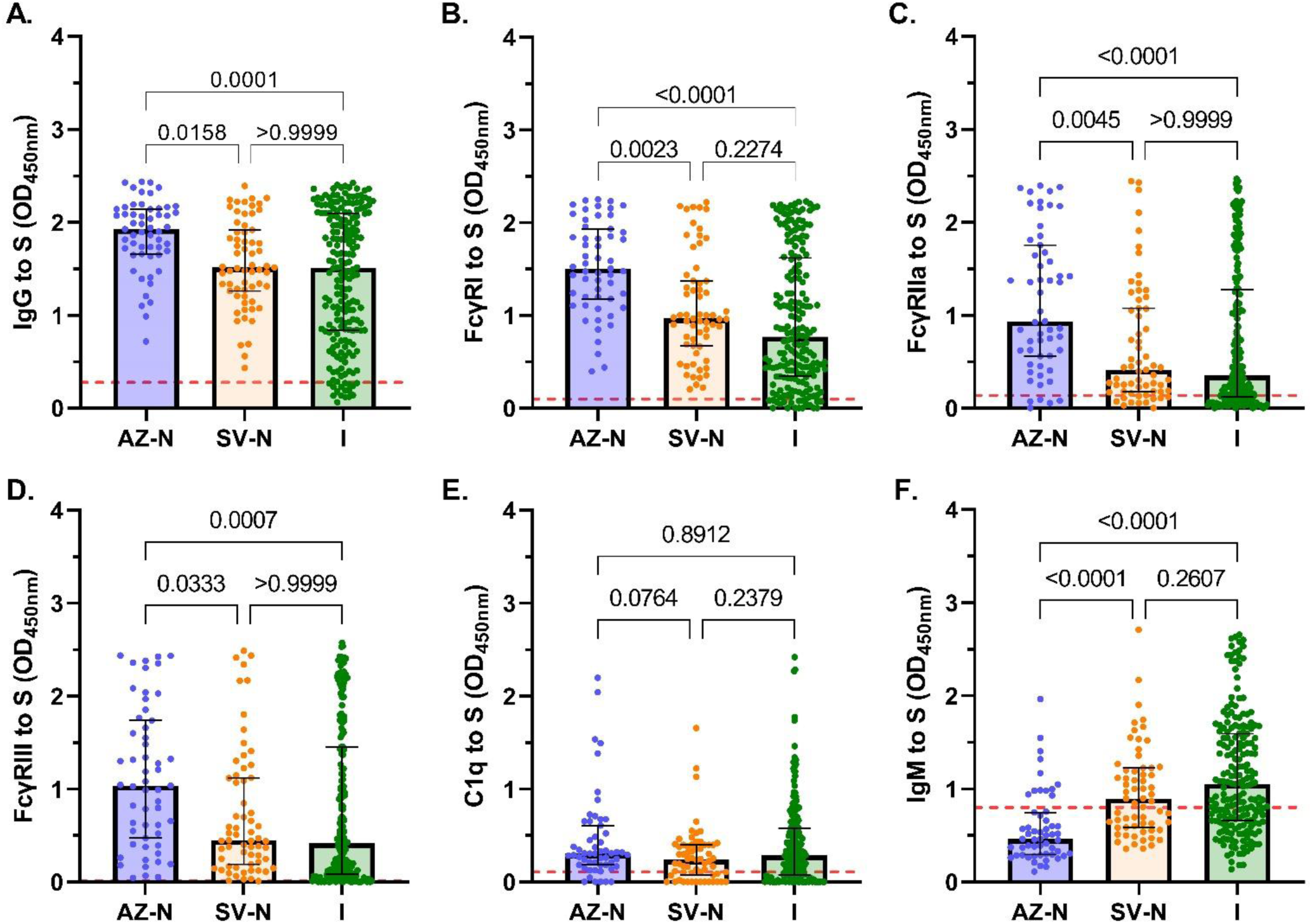
Antibody responses following AZ vaccination, SV vaccination or SARS-CoV-2 infection. Sera were collected from SARS-CoV-2 naïve individuals who were vaccinated with AZ (AZ-N; blue; n=55), or SV (SV-N; orange; n=63), and unvaccinated individuals with a known infection (I; green; n=200). Samples were tested for **(A)** IgG magnitude, binding to **(B)** FcγRI, **(C)** FcγRIIa, and **(D)** FcγRIII, **(E)** C1q-fixation, and **(F)** IgM magnitude to the SARS-CoV-2 Ancestral S protein. Data are presented as optical density (OD) read at 450nm, whereby each data point represents an individual, with columns and error bars showing median and IQR. The red dotted line indicates the seropositivity cutoff (mean + 2 SD of negative controls). Data were subjected to a Kruskal Wallis test followed by a Dunn’s multiple comparisons test.

It is noteworthy that the measure of FcγR-dimer binding agreed strongly with the activation of cells that express FcγRIIa or FcγRIII *in vitro* (P<0.0001 for both tests; **Figure 2A, B**). We evaluated the ability of serum from vaccinated individuals to activate cells expressing FcγRIIa or FcγRIII *in vitro* using a random selection of individuals with high (n=20) or low (n=20) FcγR-dimer binding activity. Serum samples were used to opsonise A549 cells expressing full-length S protein, which were then co-incubated with IIA1.6 reporter expressing cells with FcγRIIa or FcγRIII and NF-κB-nanoluciferase. There was strong agreement between the FcγR-dimer binding and cellular activation activity of serum samples.

**Figure 2.**
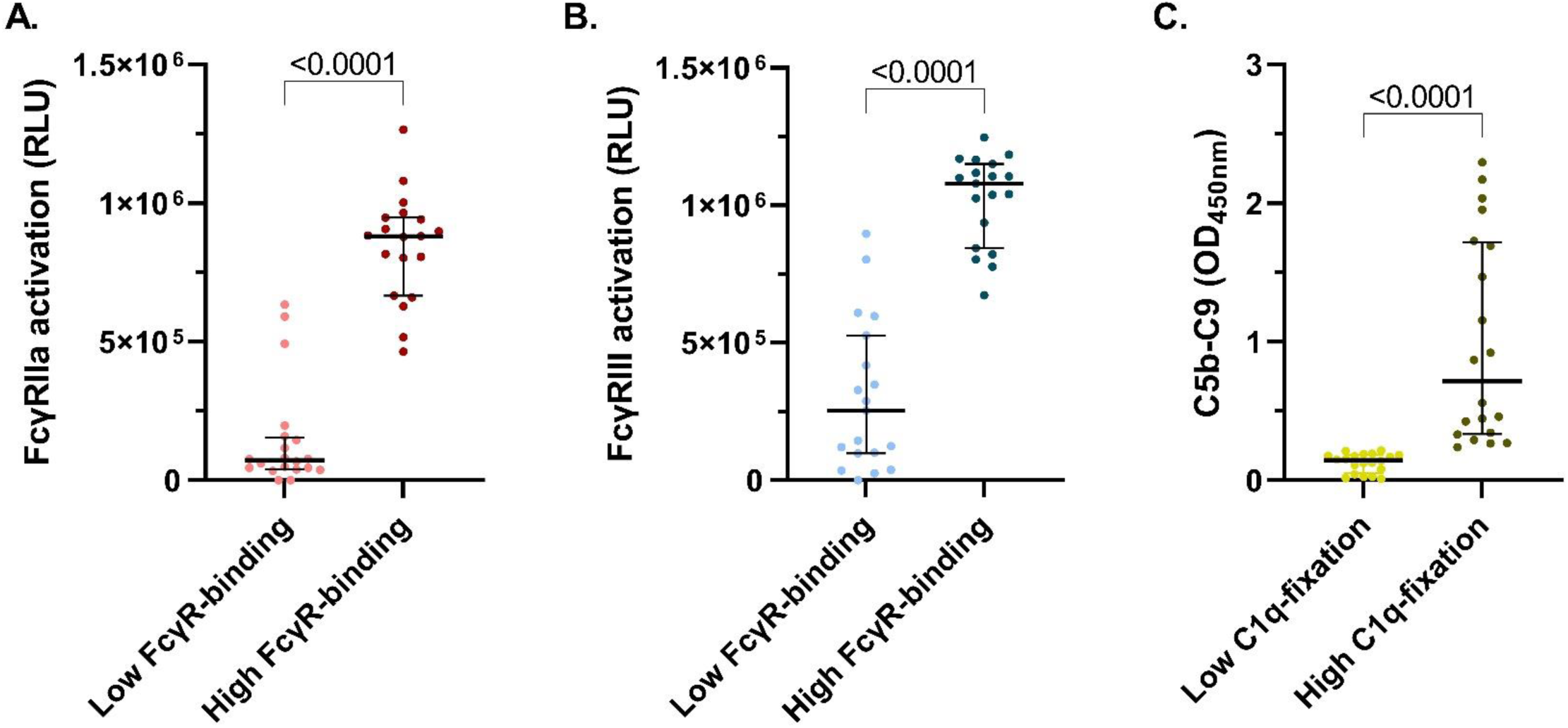
Fc-dependent cellular activation and complement activation. Serum samples from vaccinees with low (n=20) or high (n=20) FcγRIIa- and FcγRIII-dimer binding, or IgG responses were randomly selected and tested for **(A)** FcγRIIa and **(B)** FcγRIII activation using a cell-based nano-luciferase assay against A549S cells and **(C)** C5b-C9 deposition against the S protein. Dot plots of samples are shown with lines and error bars showing median ± IQR. Data are presented as optical density (OD) read at 450nm or as relative luminescence units (RLU) as indicated. Data were subjected to a Mann-Whitney U test.

Antibodies can also fix serum C1q and activate the classical complement pathway, leading to enhanced neutralisation, opsonisation, and/or lysis of pathogens. To confirm that C1q-fixation leads to downstream complement activation, we evaluated serum samples from a random selection of individuals with high (n=20) and low (n=20) IgG magnitude, for the ability to fix C1q to S protein and activate downstream complement proteins such as the terminal C5b-C9 complex. C1q-fixation activity corresponded strongly with the activation of C5b-C9 among this subset of samples (P<0.0001; **Figure 2C**). Among the AZ, SV, and infection groups, we found relatively low and comparable C1q-fixation activity, despite there being significant differences in IgG magnitude between the groups (**Figure 1E**). Given that IgM is a potent mediator of C1q-fixation (44), we quantified IgM magnitude. Interestingly, IgM was significantly higher in those vaccinated with SV compared to AZ (P<0.0001) (**Figure 1F**). Only 12 AZ vaccinated individuals had IgM levels greater than the seropositivity cutoff, whereas the majority of SV vaccinated individuals had reactivity above the cutoff. IgM was also significantly higher among the infection group compared to AZ vaccines.

The relative contribution of IgG in mediating C1q-fixation with and without adjusting for IgM was explored using multiple linear regression models. Among vaccinees, IgG but not IgM significantly correlated with C1q-fixation. The best model for vaccinated participants included IgG alone (coefficient, 0.475; R-squared, 0.291). In contrast, among infected individuals, IgG and IgM were significantly correlated with C1q-fixation in univariate analysis. The best model included IgG and IgM (coefficient for IgG with adjusting for IgM, 0.270; R-squared, 0.361; **Table S2**). This suggests that IgG is the main determinant of complement fixation activity in vaccinated individuals whereas IgM may contribute to complement fixation among infected individuals.

### Previous exposure to SARS-CoV-2 boosts vaccine-induced Fc functions

Our study also included individuals exposed to SARS-CoV-2 prior to vaccination with AZ or SV (AZ-E, n=32; SV-E, n=72). Exposure events were recorded at a similar time prior to sample collection in AZ and SV vaccinated individuals (median [IQR] days: AZ, 350 [207-443]; SV, 354 [244-388]; P=0.2604; **Table 1**). Individuals vaccinated with AZ who were previously exposed to SARS-CoV-2 had significantly greater magnitudes of all Fc functional antibody responses (FcγR-binding and C1q-fixation), as well as IgG and IgM, than AZ vaccinated individuals who were naïve (P<0.0001 for all tests; **Figure 3**). This was similarly observed in individuals vaccinated with SV with the exception of IgM, which was comparable among exposed and naïve vaccinees (**Figure 3**). The effect of prior exposure was most pronounced for hexameric C1q-fixation and FcγRIIa- and FcγRIII-dimer binding; clustering of IgG is required to get efficient engagement of these FcγRs and C1q. Of the previously exposed individuals, those vaccinated with AZ had significantly greater IgG and functional antibodies (FcγR-binding and C1q-fixation) than those vaccinated with SV (P<0.0001; **Figure 3A, C-F**). This was particularly pronounced for C1q-fixation whereby the AZ-E group median response was more than 4 times greater than the SV-E group (**Figure 3C**). These findings demonstrate that a combination of AZ vaccination and prior SARS-CoV-2 infection leads to much greater Fc-mediated functional activities.

**Figure 3.**
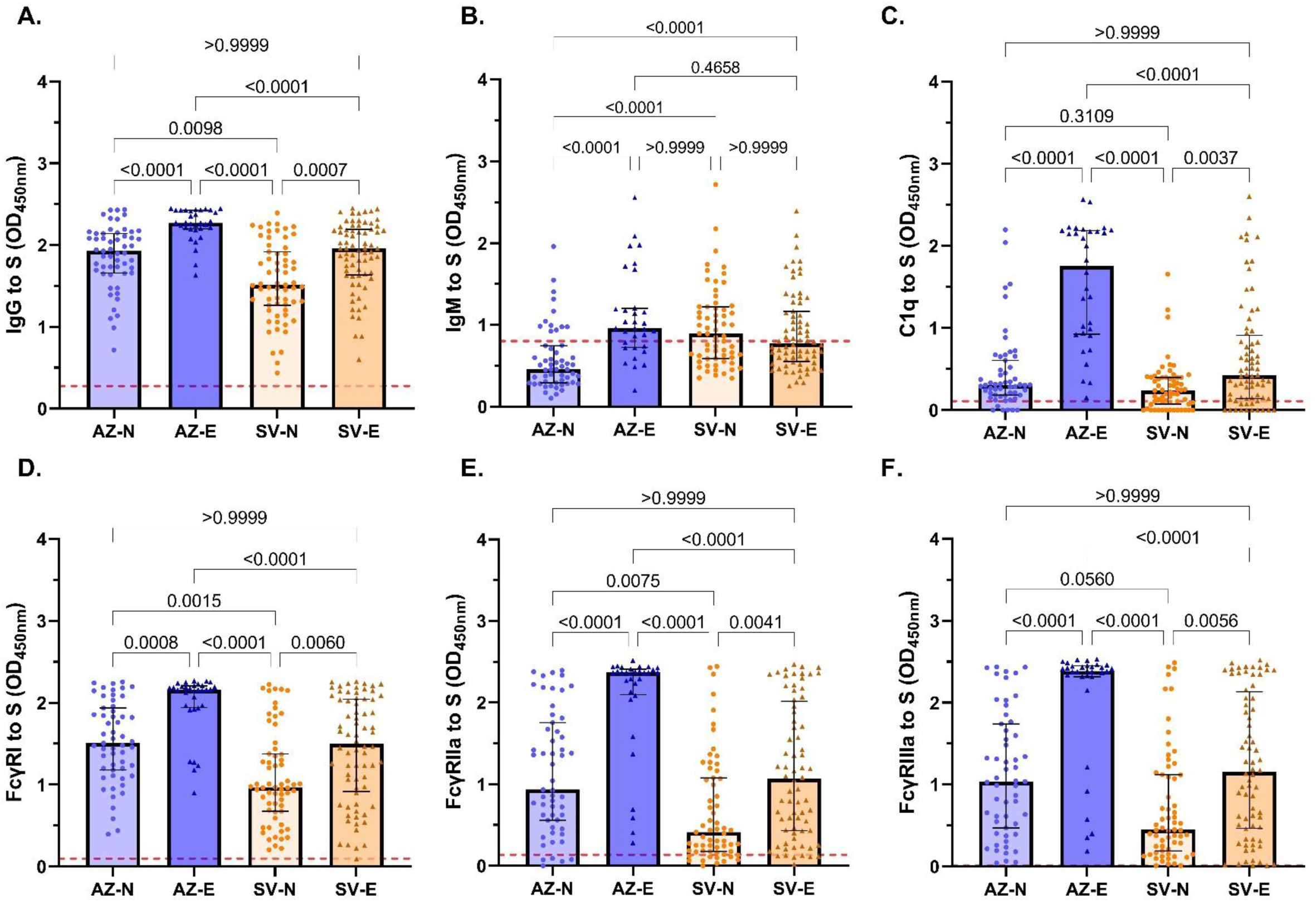
Antibody responses of infection-naïve and exposed vaccinees. Serum samples were collected from infection-naïve AZ vaccinees (AZ-N; blue; n=55), exposed AZ vaccinees (AZ-E, dark blue, n=32), infection-naïve SV vaccinees (SV-N) (orange; n=63), and exposed SV vaccinees (SV-E; brown; n=72) and tested for **(A)** IgG and **(B)** IgM magnitudes, **(C)** C1q-fixation, and **(D)** FcγRI-, **(E)** FcγRIIa-, and **(F)** FcγRIII-binding to the SARS-CoV-2 Ancestral S protein. Data are presented as optical density (OD) read at 450nm. Each data point represents an individual, with columns and error bars showing median and IQR. The red dotted line indicates the seropositivity cutoff (mean + 2 SD of negative controls). Data were subjected to a Kruskal Wallis test followed by a Dunn’s multiple comparisons test.

There were also notable differences in IgA magnitude between the groups of participants. Previously exposed AZ and SV vaccinees had greater IgA magnitude compared to their naïve vaccinee counterparts (P<0.0001 for all tests; **Figure 4**). Interestingly, the magnitude of difference between exposed and naïve vaccinees was more striking for IgA than for IgG (median OD difference: IgG 0.39; IgA 0.91). Naturally infected individuals had lower IgA magnitude than the AZ exposed (P<0.0001) and SV exposed (P=0.0025) individuals, but greater IgA magnitude compared to the SV naïve individuals (P=0.0049).

**Figure 4.**
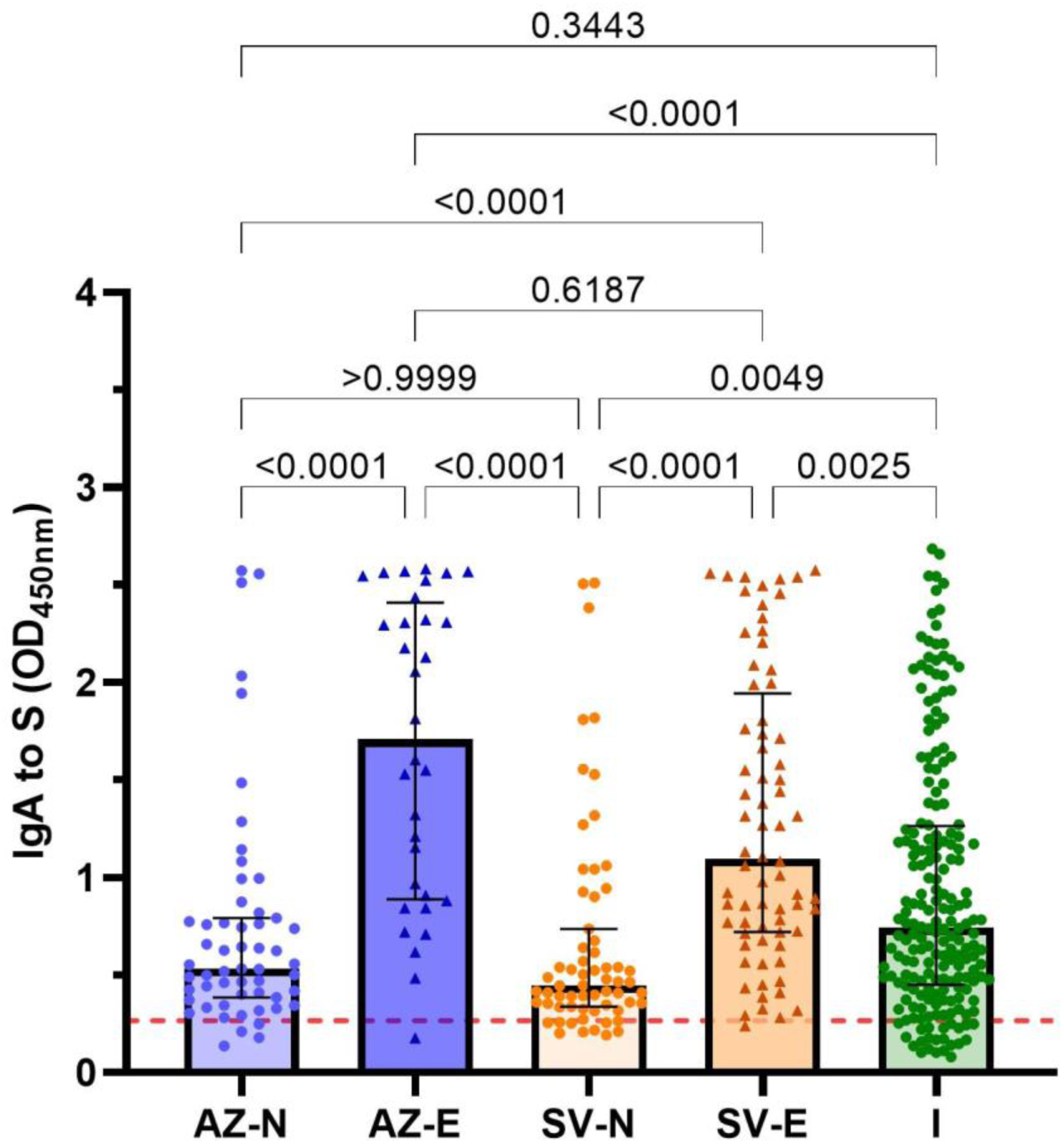
IgA magnitude of vaccinees and infected individuals. Sera were collected from infection-naïve AZ vaccinees (AZ-N; blue; n=55), exposed AZ vaccinees (AZ-E; dark blue; n=32), infection-naïve SV vaccinees (SV-N; orange; n=63), exposed SV vaccinees (SV-E; brown; n=72) and infected individuals (I; green; n=200) and tested for IgA magnitude. Data are presented as optical density (OD) read at 450nm. Each data point represents an individual, with columns and error bars showing median ± IQR. The red dotted line indicates the seropositivity cutoff (mean + 2 SD of negative controls). Data were subjected to a Kruskal Wallis test followed by a Dunn’s multiple comparisons test.

### Weak correlations between antibodies against SARS-Cov-2 S protein and age/sex

Most participants in this study were female (n=311, 73.7%); with similar proportions observed across all groups (P=0.9507; **Table 1**). There were no significant differences in antibody magnitudes and Fc-functional activities between females and males (P>0.6702 for all tests; Kruskal-Wallis test, followed by a Dunn’s multiple comparisons test). The median age of individuals vaccinated with AZ (63 years) was older compared to SV (50 years) and those with an infection (44 years) (P<0.05). Consistent with previous reports (45, 46), there was a weak correlation between age and IgG magnitude among subjects with naturally acquired antibodies in the infection group and SV-E group (r=0.2362 and r=0.3453, respectively; P<0.01 for both tests; **Table S1**). Further, age correlated weakly with Fc functional activities in the infection group (FcγRI, r=0.2059; FcγRIIa, r=0.1914; FcγRIII, r=0.2062; C1q, r=0.1714; P<0.05 for all tests) and with FcγRI and C1q in the SV-E group (r=0.2370 and r=0.2525, respectively; P<0.05 for both tests). IgG magnitude did not correlate with age amongst the naïve AZ or SV vaccinated individuals (r=0.0144 and r=-0.050, respectively; P>0.5 for both tests). However, IgM correlated negatively with age in the SV-N group (r=-0.3545, P=0.0044).

### Vaccine-induced IgG to the spike protein is retained against VOCs

The emergence of VOCs, especially the Omicron variant, continue to cause negative worldwide health impacts due to their ability to evade antibodies and immunological memory raised against the Ancestral strain through vaccination. To investigate this evasion of Ancestral specific IgG, we evaluated the ability of IgG from AZ or SV vaccinated individuals to recognise and bind to the Delta (B.1.617.2) and Omicron (B.1.1.529/BA.1) S proteins. To ensure an appropriate comparison between the ancestral, Delta, and Omicron S proteins, we confirmed the purity and integrity of all recombinant S antigens by SDS-PAGE (**Figure S1A**) and validated that the coating of each variant was comparable using the published monoclonal antibody, S2H97, that targets a conserved region at the base of the RBD (47) (**Figure S1B**). We found robust IgG reactivity to the Delta and Omicron S proteins, as was seen for the vaccine type ancestral S protein, across all vaccine groups (AZ-N, AZ-E, SV-N, and SV-E; **Figure 5**). Generally, individuals with higher anti-Ancestral S IgG levels, also had higher responses when tested against Delta and Omicron S. The magnitude of the IgG response was similar towards Ancestral and Delta S proteins in the AZ-N (P>0.9999) and SV-E groups (P>0.9999), however responses to Delta were higher in the AZ-E group (P<0.0001) and lower in the SV-N group (P=0.0099). Notably, the magnitude of the IgG response was significantly lower to Omicron compared to Ancestral S protein across all groups (P<0.0001 for all tests) except the AZ-E group (P>0.9999). High responders to the Ancestral S protein were also high responders to variant S proteins, correlating very strongly with Delta (r=0.9744) and Omicron (r=0.9306) IgG responses (P<0.0001 for both tests, Spearman’s rank correlation).

**Figure 5.**
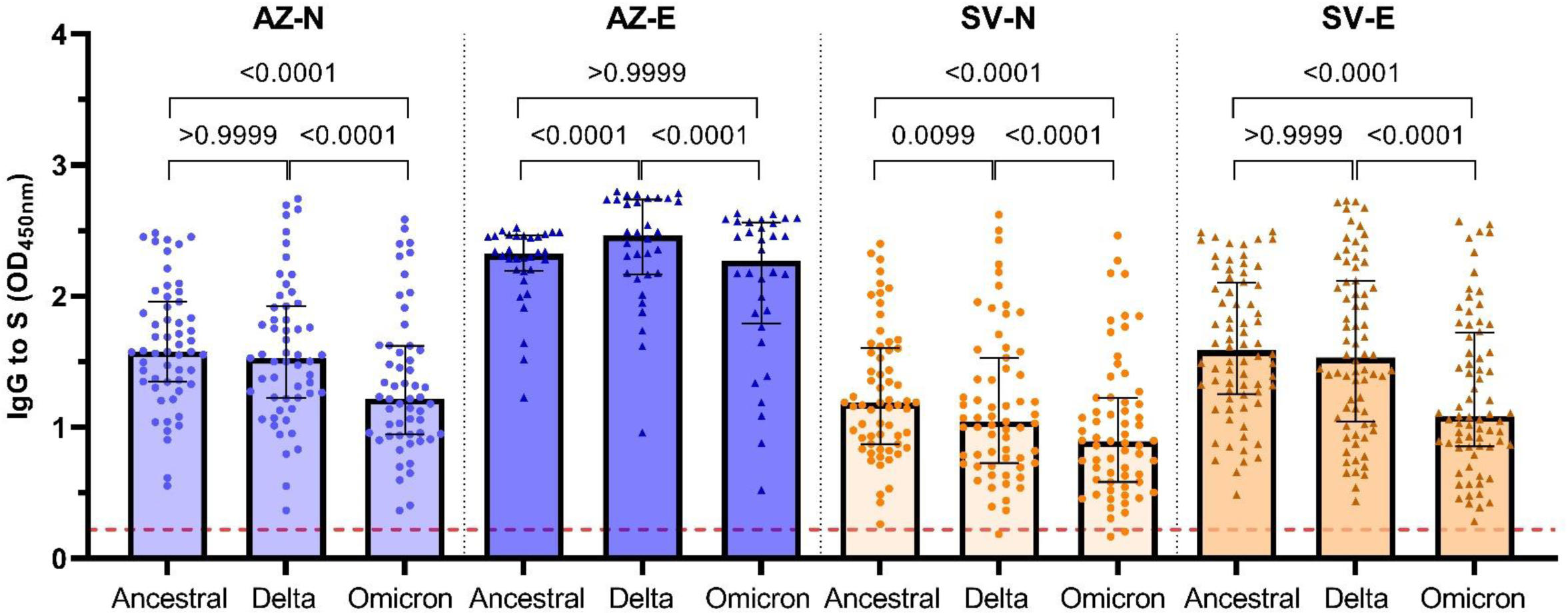
Vaccine-induced IgG to the S protein of VOCs. Sera were collected from individuals vaccinated with AZ or SV who were naïve or previously exposed to a SARS-CoV-2 infection (AZ-N, n=55; AZ-E, n=32; SV-N, n=63; SV-E, n=72). Samples were tested for IgG to the SARS-CoV-2 Ancestral, Delta, and Omicron S proteins. Data are presented as optical density (OD) read at 450nm. Each data point represents an individual and represents the average of two experiments, with columns and error bars showing median and IQR. The red dotted line indicates the seropositivity cutoff (mean + 2 SD of negative controls). Data were subjected to a Friedman test followed by a Dunn’s multiple comparisons test.

### Fc functional antibody activities are differentially retained against VOCs

All antibody FcγR-binding responses were well retained when tested against Delta, with reactivity being comparable or higher than activity to Ancestral S protein across all vaccine groups (**Figure 6A-C**). However, C1q-fixation was lower to Delta compared to Ancestral S protein in the AZ-E group (**Figure 6D**). Further, all vaccine groups displayed robust FcγRI-binding activity when tested against Omicron S, with no significant differences between Ancestral and Omicron S binding in the AZ-N and AZ-E groups. However, responses were marginally lower compared to Ancestral S in the SV-N and SV-E groups, with a median OD difference of 0.11 and 0.42, respectively (P<0.0001 for both tests; **Figure 6A**). A more pronounced loss of activity to the Omicron S protein was observed for FcγRIIa/FcγRIII-dimer binding in the AZ-N and SV-E groups. Compared to FcγRIIa-dimer binding to Ancestral S, reactivity against Omicron S was lower in the AZ-N group (0.47 OD; P=0.0013) and the SV-E group (0.56 OD; P=0.0040) (**Figure 6B**). Loss of reactivity against Omicron S was somewhat more pronounced for FcγRIII-dimer binding in the AZ-N group (0.57 OD), in the SV-N group (0.22 OD), and the SV-E group (0.75 OD) (P<0.001 for all tests; **Figure 6C**). Although several individuals in the AZ-E group had decreased activity as signified by the lowered IQR, there was no statistically significant differences in FcγRIIa- and FcγRIII-dimer binding between Ancestral and Omicron (**Figure 6B, C**). Despite this general trend of decreased FcγRIIa/FcγRIII-dimer reactivity to Omicron S across the vaccine groups, several individuals within each group maintained strong reactivity across all S proteins.

**Figure 6.**
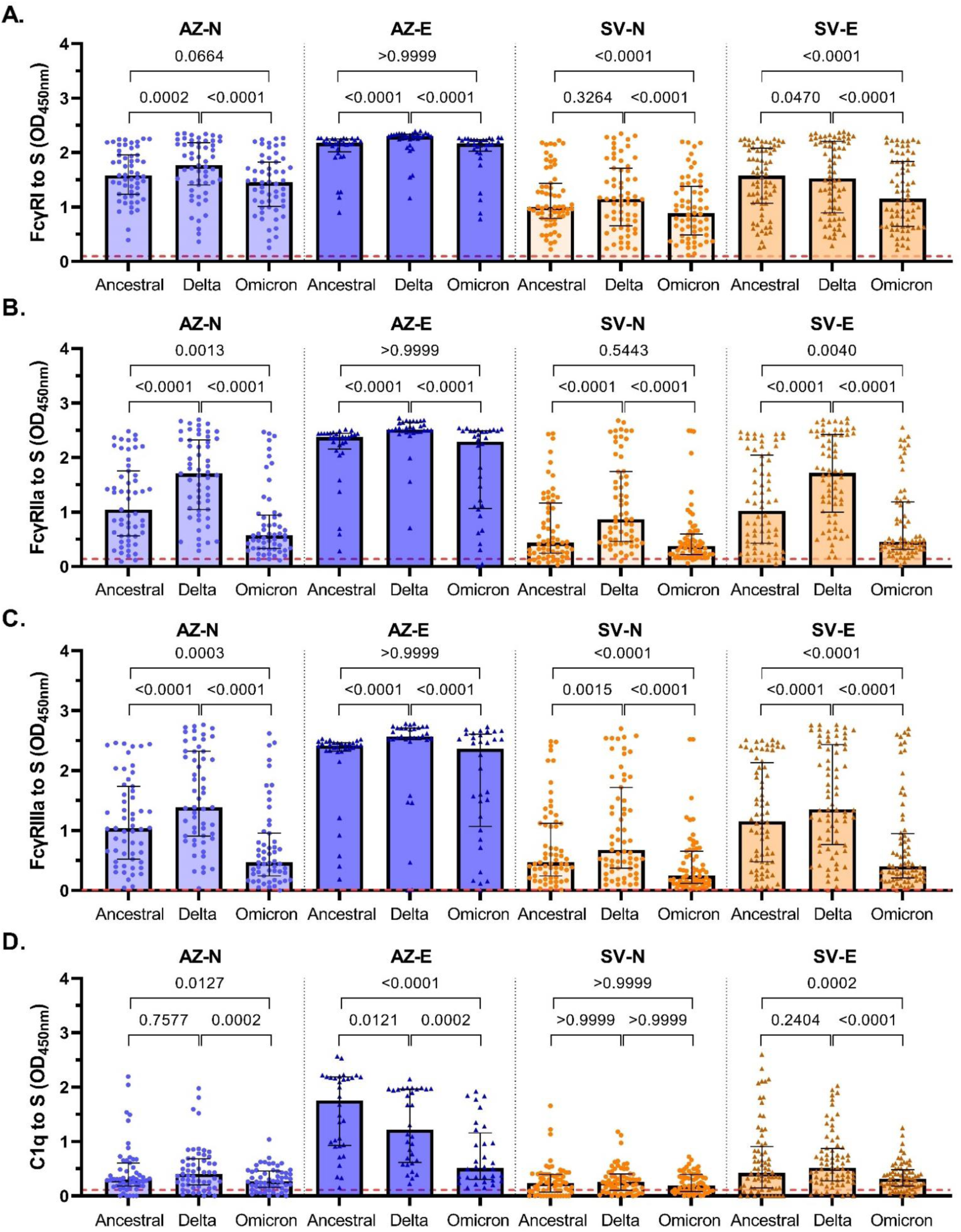
Fc functional responses against VOCs. Sera were collected from infection-naïve AZ vaccinees (AZ-N; blue; n=55), exposed AZ vaccinees (AZ-E; dark blue; n=32), infection-naïve SV vaccinees (SV-N; orange; n=63), and exposed SV vaccinees (SV-E; brown; n=72) and tested for **(A)** FcγRI, **(B)** FcγRIIa, and **(C)** FcγRIII-binding, and **(D)** C1q-fxation to the SARS-CoV-2 Ancestral, Delta, and Omicron S proteins. Data are presented as optical density (OD) read at 450nm. Each data point represents an individual, with columns and error bars showing median ± IQR. The red dotted line indicates the seropositivity cutoff (mean + 2 SD of negative controls). Data were subjected to a Friedman test followed by a Dunn’s multiple comparisons test.

Substantial C1q-fixation activity to Ancestral S was only observed among the AZ-E vaccinees. Among this group, median C1q-fixation was 3.4 times lower to Omicron S compared to Ancestral S (P<0.0001; **Figure 6D**). C1q-fixation across the AZ-N, SV-N, and SV-E groups was generally low to Ancestral and Omicron S proteins. Some individuals within each group had high reactivity to Ancestral S, which was lost when tested against Omicron S, contributing to C1q-fixation being significantly higher to Ancestral S in the AZ-N and SV-E groups (P<0.05 for both tests).

## Discussion

This study provides important new knowledge on Fc-mediated functional activities induced by two COVID-19 vaccines (AZ and SV) that have been administered to a large proportion of the world’s population. These antibody mechanisms contribute to protective immunity (24–27) but have not been extensively studied in the context of these vaccines. The Omicron variant is the dominant strain currently in circulation and has 15 mutations in the RBD compared to Ancestral RBD, which has major implications for vaccine-induced neutralising antibodies. Consequently, a marked reduction in neutralising activity of AZ and SV induced antibodies has been observed (12–14). Despite this, these vaccines retain significant efficacy against hospitalisation or death caused by Omicron (48–51), suggesting the importance of non-neutralising antibodies that can mediate function at conserved epitopes. However, the ability of AZ and SV induced antibodies to recognise and mediate function against VOCs demands further investigation, especially among diverse populations. Here, we found that vaccination with AZ induced a greater magnitude of antibodies with Fc-mediated functional activities than the SV vaccine. Vaccine responses were greater in individuals previously exposed to SARS-CoV-2 than in those who were SARS-CoV-2 naïve. This was notable, given that exposure to SARS-CoV-2 was reported on average, almost one year prior to vaccination. Encouragingly, vaccine-induced antibodies retained Fc-mediated functional activities against the Omicron S protein.

The AZ vaccine induced greater IgG and FcγR-binding responses than the SV vaccine. This difference in response magnitude was more prominent for FcγR-binding activity than for IgG which was observed in both the naïve and exposed sub-groups. This likely reflects that an opsonisation density threshold is required to mediate multivalent binding of the low affinity receptors, FcγRIIa and FcγRIII when formatted as dimers (40). IgM was only moderately induced but was significantly higher following vaccination with SV compared to AZ. Previous studies have similarly reported a low seroprevalence of anti-S IgM (52–54). Yet, robust IgM magnitudes were observed among unvaccinated individuals who were recently naturally infected with SARS-CoV-2. IgG and IgM strongly fix complement component C1q, and our analyses suggest that IgM played a more substantial role in mediating C1q-fixation amongst naturally infected individuals. Conversely, IgG was the main determinant in vaccinated individuals, as previously shown in vaccinees with low IgM (55). However, C1q-fixation responses were modest in our study, which influence the strength and significance of these associations. Therefore, larger cohorts should be evaluated to further understand the relative contribution of IgG and IgM in mediating C1q-fixation responses to SARS-CoV-2 antigens.

We observed highly variable magnitudes of Fc-mediated functional response types irrespective of the vaccine given or prior exposure status to SARS-CoV-2. Host factors may contribute to the observed variability, such as sex and age. It is noteworthy that while the majority of participants in this study were female, sex did not impact on vaccine responses as previously reported (56–58). We also found no difference in vaccine responses by age, consistent with previous reports amongst adults (59, 60); however, all participants in this study were adults aged between 22-84 years. It therefore remains unclear how the induction of Fc-dependent antibody responses may differ in children/adolescences and older individuals. Future studies are needed to further explore the determinants of vaccine responses to inform the development of more robust and efficacious vaccines.

AZ, SV, and other widely used vaccines are based on the Ancestral variant of SARS-CoV-2, so it is crucial to evaluate the functional abilities of vaccine-induced antibodies against emerging VOCs. The Delta VOC caused a large global burden of COVID-19 during 2021 but is no longer prominent. The Omicron VOC and subvariants/sublineages were responsible for the most recent epidemics, accounting for over 98% of analysed sequences worldwide since February 2022 (61). Loss of neutralising antibodies to the Omicron RBD have been widely reported, however the maintenance of real-world vaccine efficacy suggests the importance of non-neutralising functions in mediating protection from severe disease or death. In our study, FcγRI-binding was largely retained against Omicron S protein compared to Ancestral S protein, as were Fc functional responses to Delta S protein. This was likely due to vaccine-induced antibodies recognising a broad range of conserved and/or non-neutralising epitopes outside of the RBD (33, 62, 63), which were not explored in the present study.

Among the AZ-naïve and SV-exposed/naïve sub-groups, FcγRIIa- and FcγRIII-dimer binding activity was substantial, albeit lower compared to that of Ancestral S protein. FcγRI is a high affinity receptor that binds monomeric IgG, whereas FcγRIIa/FcγRIII are low affinity receptors that require immune complex formation. This partial loss of FcγRIIa- and FcγRIIa-dimer binding to Omicron, but not FcyRI-binding, likely relates to the multivalent binding of the dimeric low affinity receptors. The loss of some epitope recognition of the Omicron S, particularly in the RBD, decreases the density of opsonisation of S by IgG and this likely diminished the appropriate presentation of pairs of IgG for FcγRIIa/RIII-dimer binding. Notably, all FcγR-binding activities of antibodies against Delta and Omicron S proteins were retained among previously exposed AZ vaccinees. This was also the only sub-group with strong complement fixation activity, although this was significantly lower against Omicron. These observations indicate that a mixture of natural exposure and vaccination generated broad Fc-mediated antibody functional activities against VOCs and may partly explain the retention of real-world AZ vaccine efficacy against Omicron.

In conclusion, this study provides new insights on antibody Fc-mediated functional activities induced by the AZ and SV COVID-19 vaccines. These functions contribute to protective immunity, and a greater understanding of their induction in human populations is of great importance and relevance in the face of emerging VOCs and global pandemics. Overall, AZ was more immunogenic and induced greater Fc-mediated functional activity compared to SV, although both vaccines induced variable responses in this cohort. Notably, individuals who had received the AZ vaccine after prior SARS-CoV-2 exposure exhibited the most robust Fc-mediated activity. This finding underscores the boosting effect of vaccination on pre-existing, naturally acquired immunity. Importantly, Fc-mediated activity against the Omicron variant was relatively well retained, particularly among AZ vaccine recipients with prior infection, potentially contributing to the efficacy of AZ vaccine against Omicron VOCs. Additionally, differences between AZ and SV vaccines were more pronounced for FcγR-binding activity and complement fixation. Future research should further explore the determinants of optimal antibody responses to inform vaccine development and implementation. Overall, these insights contribute to our understanding of vaccine-induced immunity and its effectiveness against evolving variants, relevant to the prevention and amelioration of SARS-CoV-2 globally.

## Data Availability

All data produced in the present study are available upon reasonable request to the authors

## Acknowledgements and funding

We thank all study participants and staff of the UERJ health centre, Brazil. This work was funded by the National Health and Medical Research Council of Australia (Investigator Grant to JGB) and Burnet Institute. The Burnet Institute received funding from the Independent Research Institutes Infrastructure Support Scheme of the National Health and Medical Research Council of Australia, and the Operational Infrastructure Scheme of the Victorian State Government, Australia. Burnet Institute is located on the traditional land of the Boonwurrung people of the Kulin nations.

## Author Contributions

AH performed all experiments (cell activation assays were co-conducted with BW) and data analysis. JB, LCP, CV, LK, JN, and IB designed the study. AH, LK, HO, and JB led the project and analysis. HD, PP, BW, PMH, JN, and IB provided key materials and data. AH, LK, and JB wrote the manuscript with contributions from all authors.

