## Supplementary material for "Fc-dependent functional activity of ChAdOx1-S and CoronaVac vaccine-induced antibodies to the SARS-CoV-2 spike protein": Table S1, Table S2, Figure S1, Figure S2

**Table S1. Correlation of age with antibody magnitude and Fc functional activity.** Data were subjected to a Spearman’s rank correlation test (r).

|  | Age (years) Median (IQR) | **IgG** | | **IgM** | | **IgA** | | **FcγRI** | | **FcγRIIa** | | **FcγRIII** | | **C1q** | |
| --- | --- | --- | --- | --- | --- | --- | --- | --- | --- | --- | --- | --- | --- | --- | --- |
|  |  | **r** | **P** | **r** | **P** | **r** | **P** | **r** | **P** | **r** | **P** | **r** | **P** | **r** | **P** |
| **AZ-N**; n=55 | 63 (61-69) | 0.0144 | 0.9170 | -0.2441 | 0.0725 | 0.0366 | 0.7907 | 0.1220 | 0.3748 | 0.0556 | 0.6867 | 0.0212 | 0.8782 | -0.0367 | 0.7904 |
| **AZ-E**; n=32 | 62 (54-64) | 0.0766 | 0.6770 | -0.3320 | 0.0634 | 0.0738 | 0.6881 | 0.1403 | 0.4439 | 0.2560 | 0.1572 | 0.0048 | 0.9793 | 0.0961 | 0.6009 |
| **SV-N**; n=63 | 50 (40-59) | -0.0505 | 0.6944 | -0.3545 | **0.0044** | -0.0626 | 0.6261 | -0.0600 | 0.6406 | -0.1461 | 0.2532 | -0.0813 | 0.5264 | 0.1107 | 0.3879 |
| **SV-E**; n=72 | 45 (33-57) | 0.3453 | **0.0030** | -0.0233 | 0.8458 | 0.1078 | 0.3676 | 0.2370 | **0.0450** | 0.1871 | 0.1155 | 0.1834 | 0.1232 | 0.2525 | **0.0324** |
| **I**; n=200 | 44 (36-51) | 0.2362 | **0.0008** | 0.0234 | 0.7418 | 0.1383 | 0.0509 | 0.2059 | **0.0034** | 0.1914 | **0.0066** | 0.2062 | **0.0034** | 0.1714 | **0.0152** |

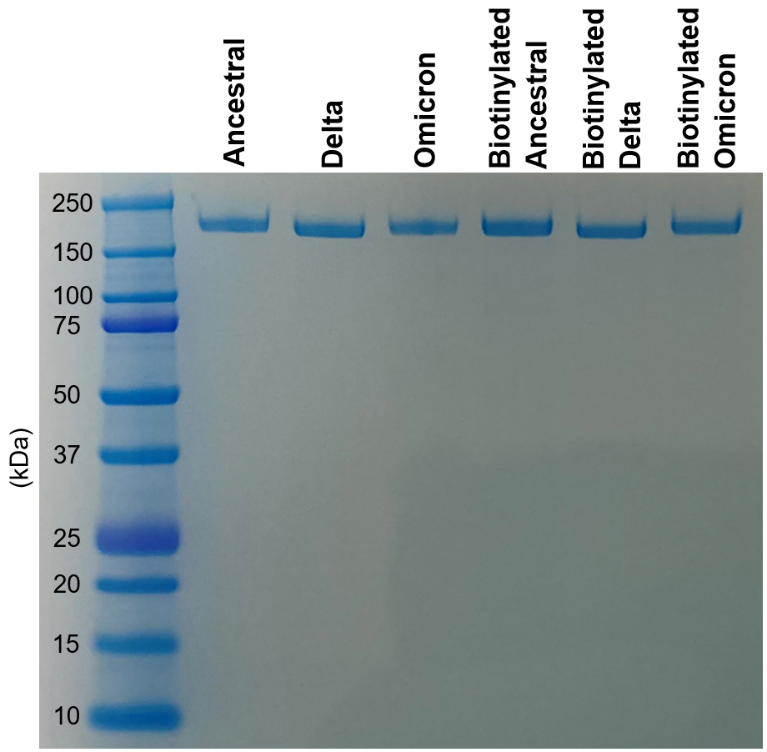

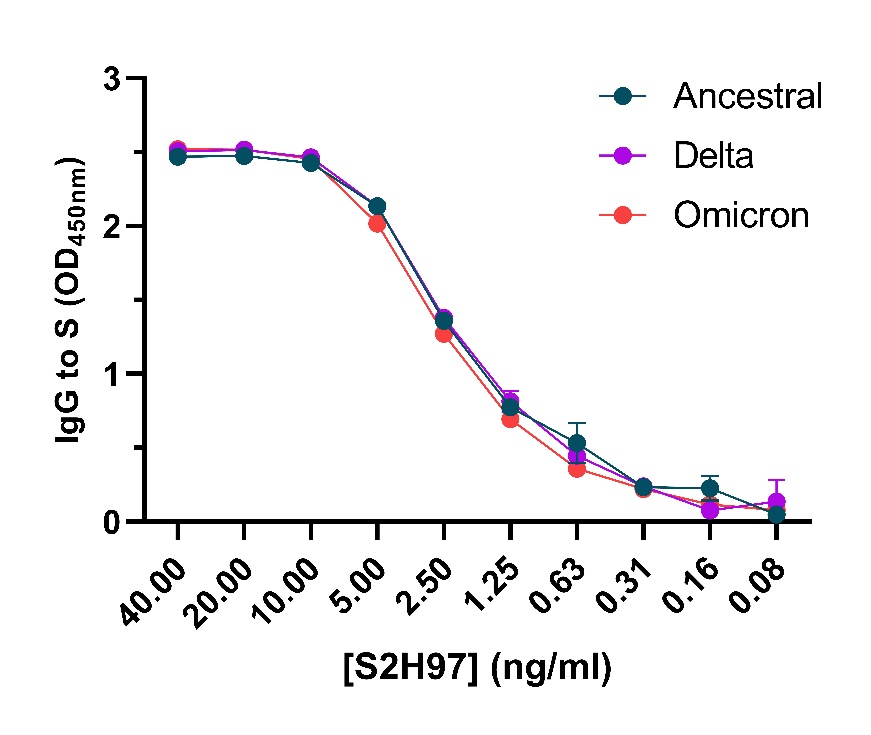

**Figure S1. Retention of IgG magnitude of the S2H97 mAb to VOCs and gel of spike antigen purity.** **(A)** The non-biotinylated and biotinylated Ancestral, Delta, and Omicron S proteins were run on a gel using LDS-NuPAGE under reducing conditions. **(B)** The S2H97 mAb was tested for IgG reactivity to the SARS-CoV-2 Ancestral, Delta, and Omicron S proteins. Data are presented as optical density (OD) read at 450nm. S2H97 mAb titration was performed in duplicate and mean ± SD are shown.

**A.**

**B.**

**Table S2. Multiple linear regression analysis of the relative contribution of IgG and IgM to C1q-fixation**

| **Participants** | **Ig Isotype** | **Coefficient** | **SE** | **95% CI** | **p-value** | **R-squared** |
| --- | --- | --- | --- | --- | --- | --- |
| **Vaccinated naïve (n=118)** | IgG | 0.475 | 0.068 | 0.341, 0.610 | <0.001 | 0.291 |
|  | IgM | 0.054 | 0.077 | -0.098, 0.208 | 0.484 | -0.004 |
|  | IgG adjusted for IgM | 0.474 | 0.068 | 0.338, 0.609 | <0.001 | 0.285 |
| **Infected (n=200)** | IgG | 0.343 | 0.037 | 0.271, 0.416 | <0.001 | 0.307 |
|  | IgM | 0.327 | 0.044 | 0.239, 0.414 | <0.001 | 0.210 |
|  | IgG adjusted for IgM | 0.270 | 0.039 | 0.193, 0.346 | <0.001 | 0.361 |

**Figure S2. Correlation matrix of IgG, IgM, and IgA magnitudes, FcγRI-, FcγRIIa-, and FcγRIII-binding, and C1q-fixation against the Ancestral S protein of individuals by sub cohorts.** Data were subjected to a Spearman’s rank correlation test (r). P values are shown in grey.

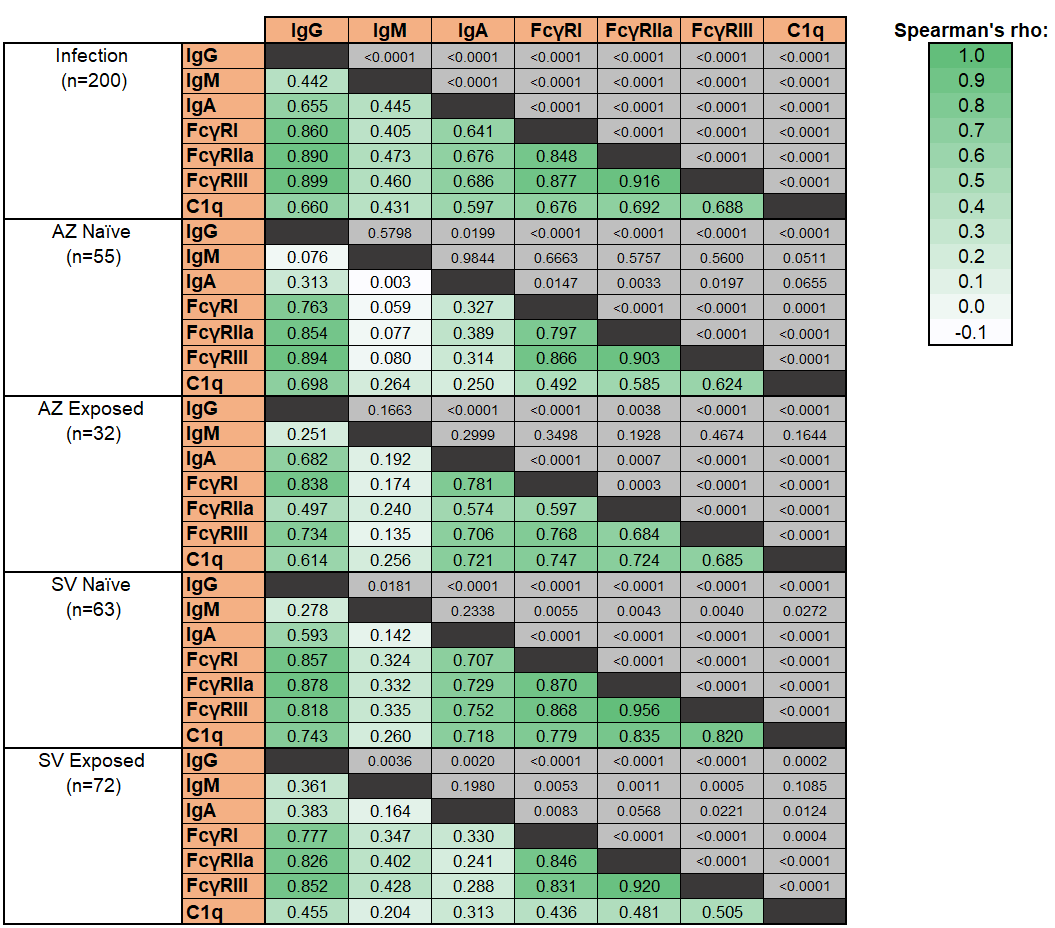
